# The effect of screening on the health burden of chlamydia: An evaluation of compartmental models based on person-days of infection

**DOI:** 10.1101/2023.06.01.23290831

**Authors:** Jack Farrell, Owen Spolyar, Scott Greenhalgh

## Abstract

Sexually transmitted diseases are detrimental to the health and economic well-being of society. Consequently, predicting impending outbreaks and identifying effective disease interventions through the use of epidemiological tools, such as compartmental models, is of the utmost importance. Unfortunately, traditional compartmental models, specifically the differential equation models attributed to the work of Kermack and McKendrick, require either a duration of infection that follows the exponential or Erlang distribution, despite the biological invalidity of such assumptions. As these assumptions negatively impact the quality of model predictions, alternative approaches are required that capture the variability in the duration of infection, along with its associated effects on the trajectory of disease, and in the evaluation of disease interventions. So, we apply an entirely new family of differential equation compartmental models based on the quantity, “person-days of infection,” to predict the trajectory of a disease. Importantly, this new family of models can alternative duration of infection distributions. As proof of concept, we calibrate our model to recent trends of chlamydia incidence in the United States and utilize a general statistical distribution that features periodic hazard rates. We then evaluate how increasing sexually transmitted disease screening rates alter predictions of incidence and saves disability adjusted life-years over a 5-year horizon. Our findings illustrate that increasing the annual screening rate of chlamydia from 35% to 40%-70% would annually avert 6.1-40.3 incidence and 1.68-11.14 disability adjusted life-years per 1000 people. This suggests increasing the screening rate of sexually transmitted diseases in the United States would greatly aid in ongoing public health efforts to curtail the rising trends in preventable sexually transmitted diseases.

## 1. Introduction

Sexually transmitted infections (STIs) have seen sharp climbs in incidence, with a ∼30% increase in the U.S. [1] between 2015 and 2019. This trend will likely be further exacerbated due to the COVID-19 pandemic, as evidence mounts on the social effects of lockdowns, the reduction in STI testing over the pandemic [2], and the diversion of health resources to more pressing matters [3]. While all STIs are of concern, chlamydia, in particular, represents a substantial health risk for the U.S., as it reigns as the most common STI, at a staggering 1.8 million incidences [4]. In part, the reason for the high incidence is the ease in which it spreads, as transmission commonly occurs through vaginal, anal, or oral intercourse, and possible mother-to-child transmission during childbirth [5]. Chlamydia is also associated with myriad negative health outcomes, including infertility [6], lymphogranuloma venereum [7], conjunctivitis [8], an increased risk of acquiring HIV [9], and social stigma [10], among numerous others. Consequently, action is required to stop the rising incidence of chlamydia, particularly through the effective deployment of health interventions.

To mitigate the spread of chlamydia, health authorities recommend several health policies. The simplest is recommending abstinence from sexual intercourse to younger demographics [5], in addition to practicing safe sex for all sexually active individuals. Health authorities also recommend that at-risk groups, namely women, gay, and bisexual men younger than 25 years [5], annually screen for chlamydia, especially for those with multiple sexual partners [5]. To communicate these recommendations, awareness campaigns on STIs are periodically launched targeting these demographics to illustrate the impact of STIs on life, reduce STI-related stigma, and to ensure people acquire the tools and knowledge to prevent and test for infection [11]. Fortunately, even if these health interventions fail and chlamydia infection occurs, effective antibiotics treatments are available, such as the use of azithromycin [12] or doxycycline [13,14]. Unfortunately, due to the delays in the appearance of symptoms and seeking of treatment, an infection can negatively affect reproductive health in both men and women. Pregnant women in particular face severe risk, as chlamydia infection may cause a fatal ectopic pregnancy, or even permanent damage to their reproductive systems through pelvic inflammatory disease (PID) [5].

The recent uptick of chlamydia incidence in the U.S. calls to light an urgent need to evaluate strategies that may curtail the trend. To inform on such strategies, we evaluate the role that screening may have in reducing chlamydia incidence, through their associated effect on symptomatic and asymptomatic durations of infection. Specifically, we developed a mathematical model that predicts chlamydia infection among the sexually active population in the U.S. The model is based on describing the trajectory of person-days of infection, rather than incidence or prevalence, due to its capacity to account for a broader variety of duration of infection distributions, while maintaining a formulation as a system of ordinary differential equations. We use this model to measure how changes in the shape of the duration of infection distribution, attributed to enhanced screening practices, affect predictions on incidence averted and disability adjusted life-years (DALYs) saved over a 5-year horizon.

### 2. Materials and methods

In what follows, we detail our mathematical model of chlamydia transmission, as characterized by a system of ordinary differential equations (ODEs). The model describes the evolution of the quantity of “person-days of infection” [15,16], which is based on the multiplication of incidence and a time-varying average duration of infection. So, we also provide details on the formulation of the time-varying average durations of infection, i.e., the mean residual waiting-times of infection, in addition to model parameters, goodness of model fit to data, the calculation of incidence averted, and DALYs averted for each intervention scenario.

### 2.1 Mathematical model

We developed a compartmental model to predict the spread of chlamydial infection across the population of the U.S. The model considers five main compartments. Each compartment has two components, the number of people and the duration. Thus, we have the person-days susceptible to infection, *Sm*, person-days latently infected, *Em*, person-days asymptotically infectious, *I*_*A*_*m*_*A*_, person-days symptomatically infectious, *I*_*SmS*_, and person-days removed from infection, *Rm*, where *m* is the average duration of chlamydia infection, *m*_*S*_ is the mean residual waiting-time of symptomatic chlamydia infection at time *t*, and *m*_*A*_ is the mean residual waiting-time of asymptomatic chlamydia infection at time *t*. The rates of transition between each compartment are given by,

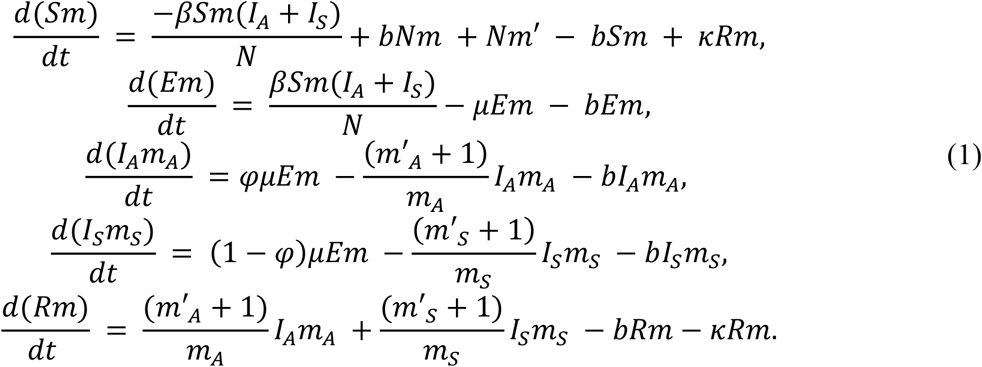

Here, *b* is the per capita birth rate, *β* is the transmission rate, *N* is the sexually active population of the U.S., 1/*μ* is the incubation period of chlamydial infection, and 1/*κ* represents the average duration of immunity to chlamydial infection. Additionally, the time-varying average duration of infection is calculated by

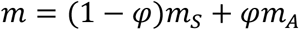

where *φ* is the proportion of asymptomatic incidence.

For our model, we consider different functional forms of mean residual waiting-times. First, we assume the classical scenario when the duration of infection is exponentially distributed, which results in a constant mean residual waiting-time. Next, we consider a generalization of a family of distributions with periodic hazard rates [17] (Supplementary Materials).

### 2.2 Parameter estimation and the durations of infection

For our model, we estimate parameters through the literature (Table 1) and published data on chlamydia incidence [18]. We estimate the average duration of asymptomatic infection with chlamydia using synthesis of data on the duration of asymptomatic chlamydia trachomatis infection [19,20] (Table 1). Additional model parameters are estimated using a nonlinear least squares procedure, in conjunction with Matlab’s ode45 and fmincon algorithms, which fit the SEAIR and gSEAIR models to weekly chlamydia incidence (Figure 2). Additional parameter details, including those for the calculation of DALYs, are available in Table 1 and the Supplementary Materials.

**Table 1.** Parameters, values, and sources.

| Symbol | Parameter | Base value | Source |
| --- | --- | --- | --- |
| $N$ | Sexually active population | 15.5 million | Fit from data |
| $b$ | Birth rate/death rate | 0.024/year | [36] |
| $\beta$ | Transmission rate | 0.050 – 0.062/year | Table S.1. |
| $1/\kappa$ | Avg. duration of immunity | 90 days | [22] |
| $1/\mu$ | Incubation period | 14 days | [22] |
| $\varphi$ | Proportion of asymptomatic infections | 0.77 | [37] |
| $\mu_A$ | Avg. duration of asymptomatic infection | 190 days | [19] |
| $\mu_S$ | Avg. duration of symptomatic infection | 10 days | [38] |
| $\omega_I$ | Disability weight of symptomatic infection | 0.006 | [39] |
| $\omega_A$ | Disability weight of asymptomatic infection | 0 | [39] |
| $\omega_M$ | Disability weight of moderate pelvic inflammatory disease | 0.114 | [39] |
| $\omega_D$ | Disability weight of severe pelvic inflammatory disease | 0.324 | [39] |
| $\omega_E$ | Disability weight of epididymo-orchitis | 0.128 | [39] |
| $\rho_I$ | Proportion of symptomatic incidence with no complications | 0.23 | [37] |
| $\rho_A$ | Proportion of asymptomatic incidence with no complications | 0.60 | [37] |
| $\rho_M$ | Proportion of incidence leading to PID | 0.09 | [38] |
| $\rho_E$ | Proportion of incidence with epididymo-orchitis | 0.0415 | [40] |
| $\rho_D$ | Proportion of PID cases classified as severe | 0.175 | Supplementary Materials |
| $\rho_{Death}$ | Proportion of PID cases resulting in death | $2.9 \times 10^{-6}$ | [39] |
| $\lambda_I$ | Avg. duration of uncomplicated symptomatic infection | 0.027 years | [38] |
| $\lambda_A$ | Avg. duration of uncomplicated asymptomatic infection | 0.521 years | [19] |
| $\lambda_M$ | Avg. duration of moderate PID due to infection | 21.01 years | Supplementary Materials |
| $\lambda_D$ | Avg. duration of severe PID due to infection | 21.01 years | Supplementary Materials |
| $\lambda_E$ | Average duration of epididymo-orchitis and its complications due to infection | 25.38 years | Supplementary Materials |
| $\lambda_{Death}$ | Avg. duration of life lost from death due to PID | 36.5 years | Supplementary Materials |
| $\gamma_{RCF}$ | Avg. time period of reproductive capability in women | 39 years | Supplementary |

|  |  |  | Materials |
| --- | --- | --- | --- |
| $\gamma_{RCM}$ | Avg. time period of optimal reproductive capability in men | 41.6 years | Supplementary Materials |
| $\varrho_F$ | Avg. time period females are able to reproduce before contracting chlamydial infection | 8.5 years | Supplementary Materials |
| $\varrho_M$ | Avg. time period males are able to reproduce before contracting chlamydial infection | 8.6 years | Supplementary Materials |
| $\vartheta_{FP}$ | Years lost in reproductive capability in women due to PID | 9.49 years | Supplementary Materials |
| $\vartheta_{ME}$ | Years lost in optimal reproductive capability in men due to epididymo-orchitis | 7.62 years | Supplementary Materials |
| $\tau$ | Proportion of chlamydial infections held by women | 0.6624 | [41] |
| $\omega_I$ | Disability weight of symptomatic chlamydia with no complications | 0.006 | [39] |
| $\omega_A$ | Disability weight of asymptomatic chlamydia with no complications | 0.0 | [39] |
| $\omega_M$ | Disability weight of incidence leading to PID | 0.114 | [39] |
| $\omega_E$ | Disability weight of incidence with epididymo-orchitis | 0.128 | [39] |
| $\omega_D$ | Disability weight of PID cases classified as severe | 0.324 | [39] |

**Figure 1.**
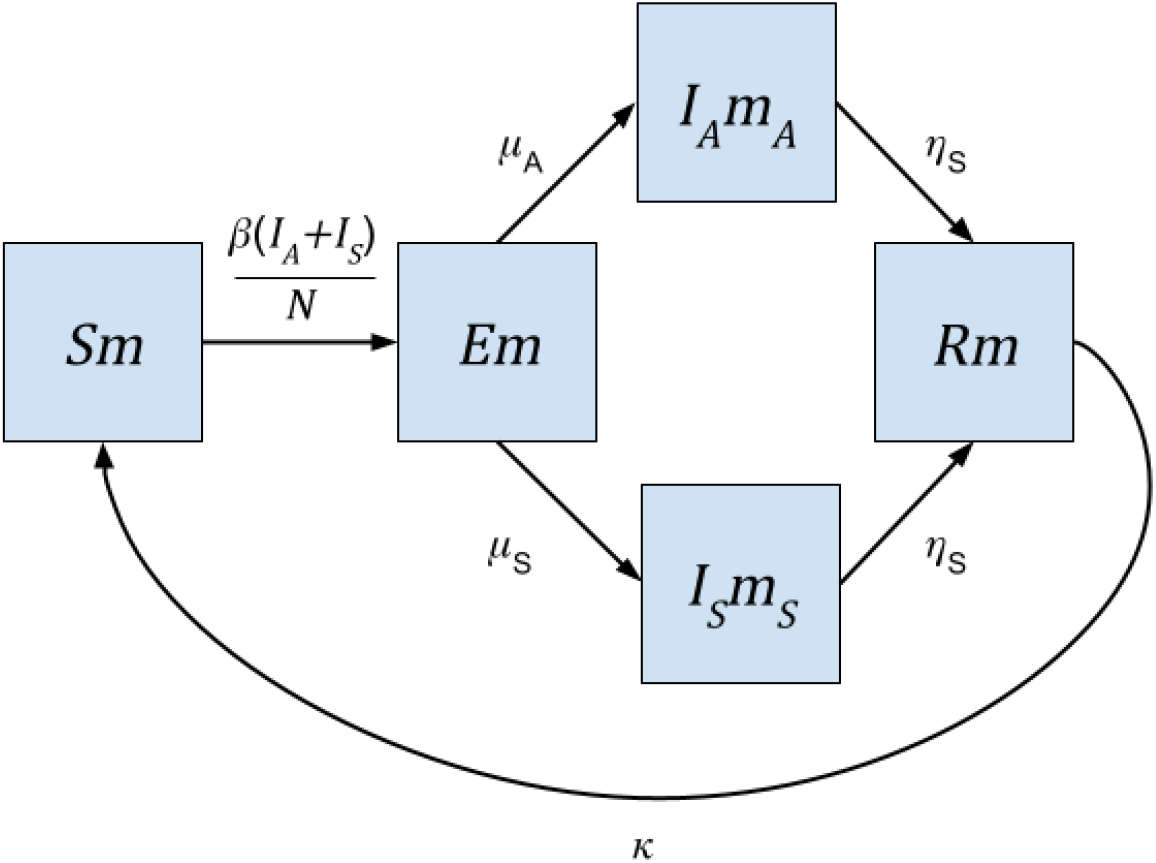
Compartmental diagram. The flow of susceptible person-days (*Sm*), to latently infected (*Em*), and either asymptomatically infectious person-days (*I*_*A*_*m*_*A*_), or symptomatically infectious person-days (*I*_*S*_*m*_*S*_), and recovered person days (*Rm*). Compartments are composed of individuals multiplied by the time-varying average duration of infection, *m*, the time-varying duration of asymptomatic infection, *m*_*A*_, or the time-varying duration of symptomatic infection, *m*_*S*_. For ease of presentation, birth and mortality rates are not included (see equation 1 for details).

**Figure 2.**
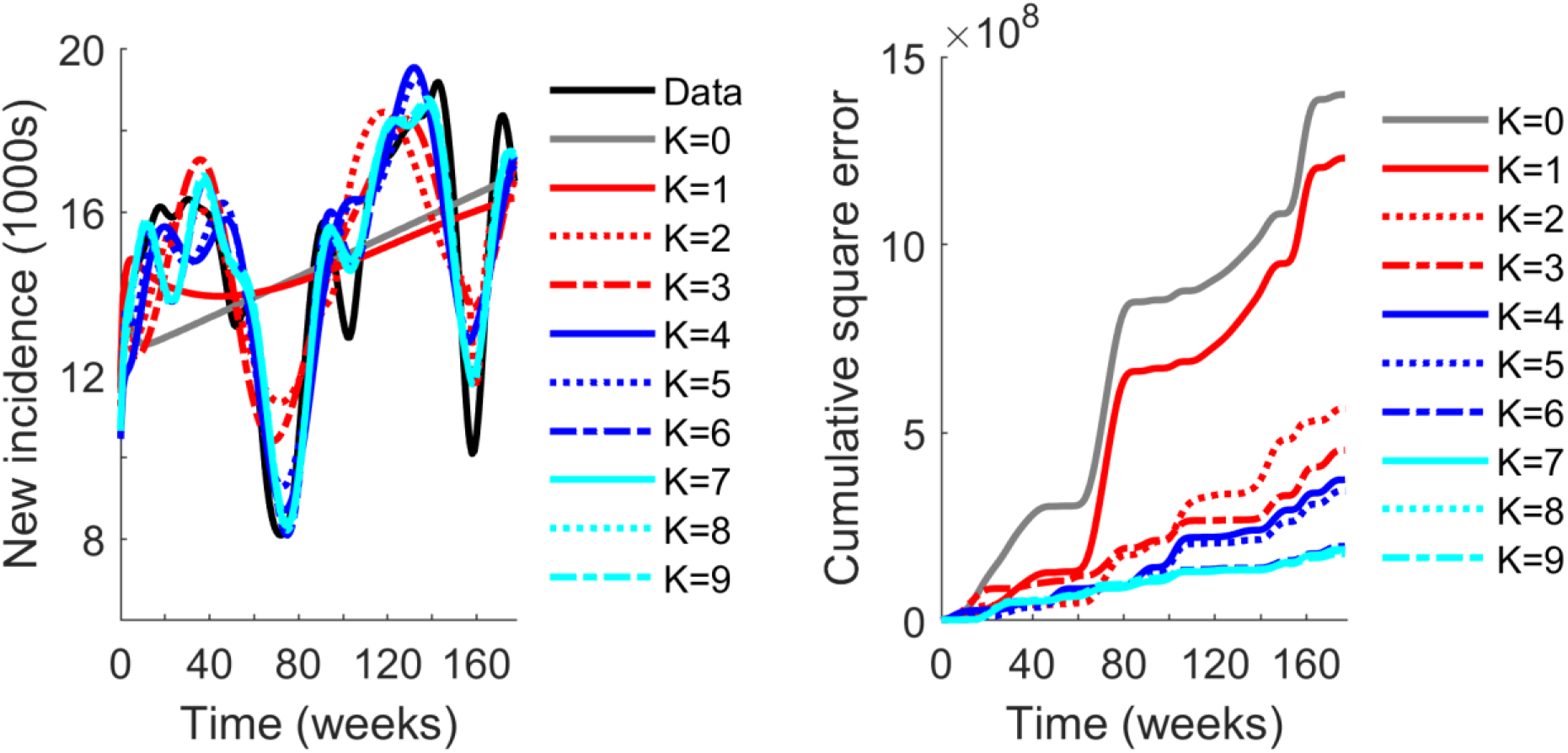
Trajectories of new chlamydia incidence and cumulative square error. a) The trajectory of chlamydial infection in the United States over 175 weeks, and b) the square error of model predictions relative to reported data. Reported incidence (black curve), a classical SEAIR model fit (*K =* 0, grey curve), and the gSEAIR models with hazard rates that feature one to nine cosine terms (*K =* 1 to *K = 9*).

Traditionally, the duration of infection is assumed to be Exponentially distributed. Under such assumptions

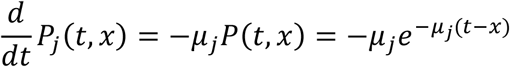

where *P*_*j*_ is a survival function, *μ*_*j*_ is the mean, and the subscript *j* is used to denote either asymptomatic (A) infection or symptomatic (S) infection [21].

Given this definition of the duration of infection distributions, the mean residual waiting-time is

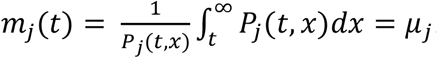

Note, because *m*_*j*_*(t)* is constant it follows that system (1) reduces exactly to the traditional SEAIR model.

To evaluate the potential periodicity of the trajectory of chlamydia, we assume both the asymptomatic and symptomatic durations of infection follow a family of distributions with periodic hazard rates. This family of distribution is based on generalizing the simplest probability density function, *p(t)*, with a periodic hazard rate [17], namely

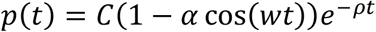

where *C* is a normalizing constant, *ρ* >0, *α* ∈ (−1,1) and *w* ∈ [0,2*π*], to that of a Fourier cosine series,

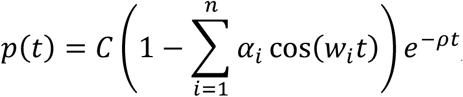

where 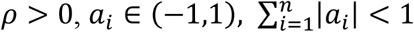 and *w*_*i*_ ∈ [0,2π].

Recalling that 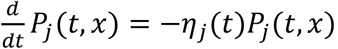, the hazard rate is given by (Supplementary Materials)

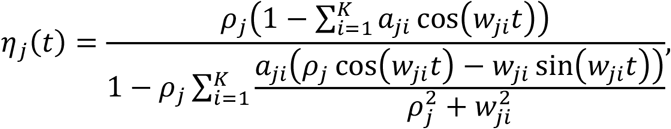

with *ρ*_*j*_ > 0, *a*_*ji*_ ∈ (−1,1), and 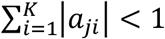. Here *ρ*_*j*_ represents the average duration of infection in the absence of periodic effects, *a*_*ji*_ is the amplitude of variation in recovery with frequency 2π/*w*_*ji*_, where the subscript *j* is used to denote either asymptomatic (A) infection or symptomatic (S) infection [21].

Given the hazard rate and its relation to the mean residual waiting-time, *ηj = (m*_*j*_*′ +* 1)/*m*_*j*_ with *m*_*j*_(0*) = μ*_*j*_, we have that

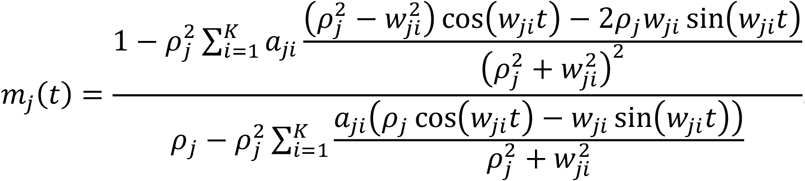

### 2.4 Intervention scenarios and health metrics

To inform on the benefit of awareness campaigns for mitigating chlamydia transmission, we consider the effects of increasing STI screening rates.

We assume that the proportion of people that get screened within one year [22] follows the distribution

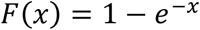

Thus, the average time a person has between screenings is 1/*x* years, which yields the screening rate [22],

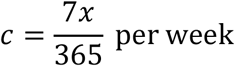

Imposing a baseline annual screening rate of 35% [23] of the population, it follows that *c*_*baseline*_ *=* 0.00*83 per week*. Alternatively, if we consider intervention scenarios that increase the annual proportion of the population screened to 40%, 50% and 70%, we have that *c*_40_ *=* 0.00*98 per week, c*_50_ *=* 0.01*33 per week*, and *c*_70_ *=* 0.0231 *per week* respectively.

For these screening rate scenarios, we evaluate our model over 5 years. We estimate incidence under each mean residual waiting-time by subtracting predictions from 40%, 50%, and 70% screening rates from the baseline. The same approach was also taken to estimate annual DALYs saved, which were discounted at the standard rate of 5% per year (see Supplementary Materials for further details).

### 2.5 Goodness of fit

To evaluate the quality of model fit to incidence data, we calculate the Akaike information criterion (AIC) [24].

For ease of presentation, we define the list of variables as *X: = (S, E, A, I, R, m*_*A*_, *m*_*S*_)^*T*^, and the list of parameters as

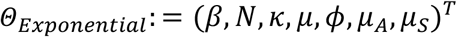

when the duration of infection is exponentially distributed, and

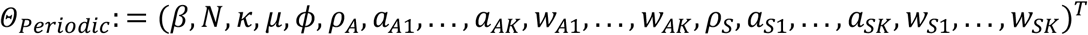

when the duration of infection follows the distribution with a periodic hazard rate.

Thus, defining *Ψ = (s*_0_, *e*_0_, *a*_0_, *i*_0_, *r*_0_, *m*_*A*_(0*), m*_*s*_(0))^*T*^, we can represent the ODE system as

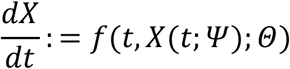

The new symptomatic infections are defined by

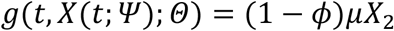

For these models the Residual Sum of Squares (RSS) is

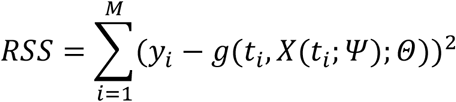

where *y*_*i*_ is the observed incidence on the *i*^*th*^ week [14,25] and *M =* 175 is the number of data points. Optimal parameters sets, 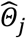, and initial conditions 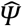 for the *j*^*th*^ distribution types were then determined by minimizing RSS (Figure 4) through a combination of Matlab’s ode45 and fmincon algorithms.

Thus, given the optimal parameters, it follows that

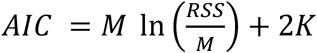

where *K* represents the number of model parameters to be estimated from observed data. The model with minimum *AIC* is deemed the best fit.

From the AIC, we approximate the probability that model *i*^*th*^ is the best candidate among all models (in the sense of combining accurate predictions while limiting the possible number of parameters[26]) by calculating AIC weights. First defining *ΔAIC*_*j*_ *= AIC*_*j*_ − min(*AIC*), the Akaike weights [27] for each scenario are

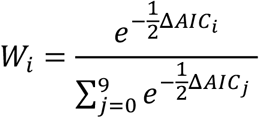

## 2. Results

We assessed the effectiveness of the gSEAIR model by estimating the health burden of chlamydia in the US and informing on the potential health benefits of increasing STI screening rates from the current annual coverage of 35% to 40%, 50%, and 70%, respectively. For these scenarios, we estimated the annual incidence averted and DALYs averted per year relative to the baseline for both the traditional SEAIR and gSEAIR models. The particular gSEAIR models considered feature mean residual waiting-times for the duration of asymptomatic and symptomatic infection with up to 9 cosine terms (*K =* 1 to *K = 9*). To identify the most appropriate SEAIR and gSEAIR models, we used AIC, in addition to AIC weights, to both identify the optimal model and estimate the probability it was optimal among all considered scenarios.

Our results show that increasing the number of cosine terms in gSEAIR decreases the square error of model predictions relative to the data (Figure 3). Additionally, the gSEAIR model based on a hazard rate with 6 cosine terms is optimal compared to the other candidate models (Figure 3), as it had the lowest AIC score (Table 3). Conversely, the SEAIR model (i.e., the gSEAIR model with *K =* 0) had the highest AIC score, with a ΔAIC of 300.6 (Table 3), which indicates it is the least effective of the modeling scenarios considered. This result is supported by the AIC weights, which indicate that the *K = 6* scenario of the gSEAIR model is optimal with a probability of 0.54 (Table 3), where most other scenarios had ΔAIC scores of at least 4.5 and AIC weights below 0.05. The exception to this is *K = 7*, where the ΔAIC was 0.81 and the AIC weight was 0.36 (Table 3), suggesting this scenario is a viable alternative to the *K = 6*.

**Table 2.** Duration of infection distribution parameters with periodic hazard rate (*K = 6*).

| Symptomatic parameters |  |  |  | Asymptomatic parameters |  |  |  |
| --- | --- | --- | --- | --- | --- | --- | --- |
| Amplitude |  | Period (days) |  | Amplitude |  | Period (days) |  |
| $a_{S1}$ | 0.19 | $2\pi/w_{S1}$ | 198.7 | $a_{A1}$ | 0.45 | $2\pi/w_{A1}$ | 817.0 |
| $a_{S2}$ | 0.22 | $2\pi/w_{S2}$ | 82.4 | $a_{A2}$ | 0.18 | $2\pi/w_{A2}$ | 97.0 |
| $a_{S3}$ | 0.19 | $2\pi/w_{S3}$ | 57.3 | $a_{A3}$ | -0.04 | $2\pi/w_{A3}$ | 56.1 |
| $a_{S4}$ | 0.21 | $2\pi/w_{S4}$ | 44.0 | $a_{A4}$ | 0.15 | $2\pi/w_{A4}$ | 45.0 |
| $a_{S5}$ | 0.02 | $2\pi/w_{S5}$ | 36.4 | $a_{A5}$ | 0.01 | $2\pi/w_{A5}$ | 36.5 |
| $a_{S6}$ | 0.10 | $2\pi/w_{S6}$ | 30.2 | $a_{A6}$ | 0.17 | $2\pi/w_{A6}$ | 30.5 |

**Table 3.** Chlamydia incidence averted and DALYS saved for screening rates that achieve 35%, 40%, 50%, and 70% coverage of the population per year.

| | $K$ | | | | | | | | | |
| --- | --- | --- | --- | --- | --- | --- | --- | --- | --- | --- |
|  | 0 | 1 | 2 | 3 | 4 | 5 | 6 | 7 | 8 | 9 |
| AIC Score<br>(1000s) | 3.333 | 3.319 | 3.318 | 3.157 | 3.131 | 3.124 | 3.033 | 3.034 | 3.037 | 3.03 |
| ΔAIC Score | 300.6 | 285.7 | 154.6 | 123.9 | 98.0 | 91.0 | 0 | 0.8 | 4.6 | 4.8 |
| AIC weights | 0 | 0 | 0 | 0 | 0 | 0 | 0.54 | 0.36 | 0.05 | 0.05 |
| Annual incidence/1000 ppl |  |  |  |  |  |  |  |  |  |  |
| Baseline ($c_{30}$) | 55.5 | 55.9 | 56.0 | 55.9 | 55.6 | 55.9 | 55.9 | 55.9 | 55.9 | 56.1 |
| Annual incidence averted (1000 ppl) |  |  |  |  |  |  |  |  |  |  |
| 40% screened<br>($c_{40}$) | 6.3 | 6.1 | 7.2 | 6.8 | 7.9 | 7.9 | 8.1 | 8.2 | 8.3 | 8.2 |
| 50% screened<br>($c_{50}$) | 17.9 | 17.5 | 19.8 | 19.1 | 21.8 | 21.5 | 22.1 | 22.3 | 22.5 | 22.4 |
| 70% screened<br>($c_{70}$) | 35.8 | 35.2 | 37.6 | 36.8 | 39.4 | 39.3 | 39.8 | 40.1 | 40.2 | 40.3 |
| DALYs saved per year (1000 ppl) |  |  |  |  |  |  |  |  |  |  |
| 40% screened<br>($c_{40}$) | 1.73 | 1.68 | 1.97 | 1.89 | 2.17 | 2.18 | 2.24 | 2.24 | 2.29 | 2.24 |
| 50% screened<br>($c_{50}$) | 4.94 | 4.83 | 5.46 | 5.28 | 6.01 | 5.93 | 6.07 | 6.15 | 6.19 | 6.18 |
| 70% screened<br>($c_{70}$) | 9.89 | 9.74 | 10.40 | 10.18 | 10.90 | 10.86 | 11.01 | 11.09 | 11.10 | 11.14 |

**Figure 3.**
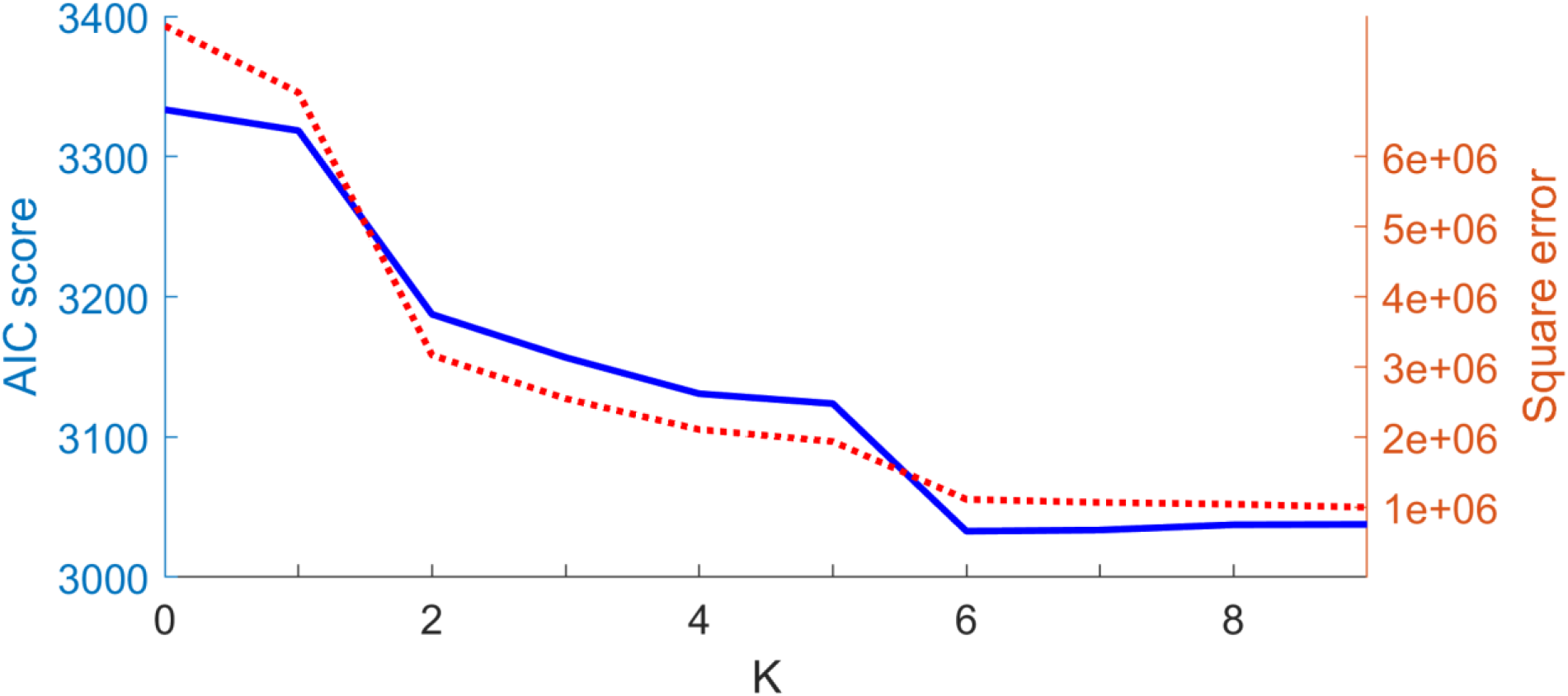
Akaike information criterion score and square error. The AIC score (solid blue curve, left axis) and square error (dotted orange curve, right axes) for SEAIR (*K =* 0) and gSEAIR (*K =* 1 to *K = 9*) models relative to incidence data.

**Figure 4.**
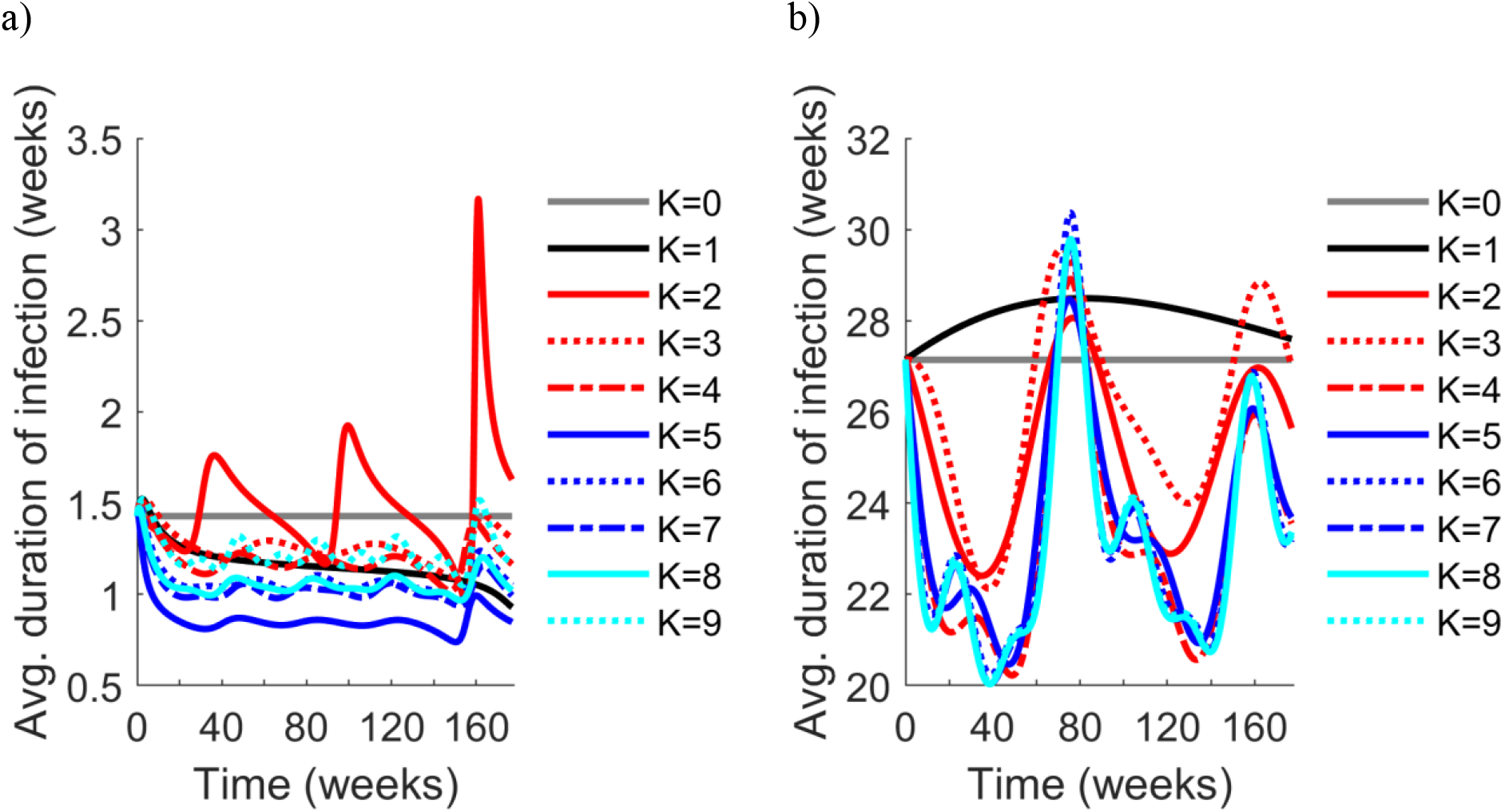
Mean residual waiting-times of the duration of infection of chlamydia. The average duration of infection over 175 weeks given a) symptomatic infection and b) asymptomatic infection. The value of *K* corresponds to the number of cosine terms in the Fourier cosine series in the probability density function, with *K =* 0 corresponding to an exponential density function.

At a baseline screening rate of 35%, the SEAIR and gSEAIR models predicted 55.5-56.1 annual incidences of chlamydia per 1000 people. Increasing the screening rate to 40%, 50%, or 70% averted 6.1-8.3, 17.5-22.3, and 35.2-40.3 annual incidences per 1000 people, respectively, depending on the number of cosine terms included in the mean residual waiting-times (Table 3). Of these findings, the gSEAIR model generally predicted a greater benefit when increasing the screening rate to 40%-70%, with an additional 1.8-4.2 annual incidence averted per 1000 people when compared to the gSEAIR model (*K* = 6) to the SEAIR model.

With regard to the health burden of chlamydia, we predict increasing the screening rate to screening rate 40%, 50%, or 70% will annually avert 1.68-2.29, 4.83-6.19, and 9.74-11.14 DALYs per 1000 people (Table 3), respectively. Typically, the gSEAIR models predicted a greater quantity of DALYs averted relative to the SEAIR model, except for the *K =* 1 case (Table 3). When comparing the optimal gSEAIR model (*K = 6*) to the SEAIR model, predictions illustrate an additional annual 0.51 DALYs averted per 1000 people (Table 3). Averaging across all scenarios, annual DALYs averted per 1000 people were 2.06, 5.7, and 10.6 for screening rates of 40%, 50%, and 70%, respectively (Table 3).

Averaging all gSEAIR modeling scenarios illustrates the duration of asymptomatic infection peaked at 32.2 weeks around the 85^th^ week of the outbreak. The symptomatic infection peaked much earlier, specifically around week 22, with an average duration of 1.9 weeks (Figure 4). Interestingly, the only mean residual waiting-time (with *t* >0) that was strictly greater than the constant average duration of asymptomatic infection for the SEAIR model was the gSEAIR with *K =* 1. In contrast, mean residual waiting-times were typically less than the average duration of symptomatic infection for the SEAIR model, although several cases briefly surpass this value near the end of the outbreak (Figure 4). Towards this regard, when *K = 5*, the mean residual waiting-time for symptomatic infection was at a minimum (Figure 4), although this may be a result of the associated probability density function decay rate (Figure 5). For asymptomatic infection, there is not a clear scenario where one of the mean residual waiting-times is consistently the minimum, as the majority of scenarios appear to converge to a common probability density function (Figure 4, Figure 5).

**Figure 5.**
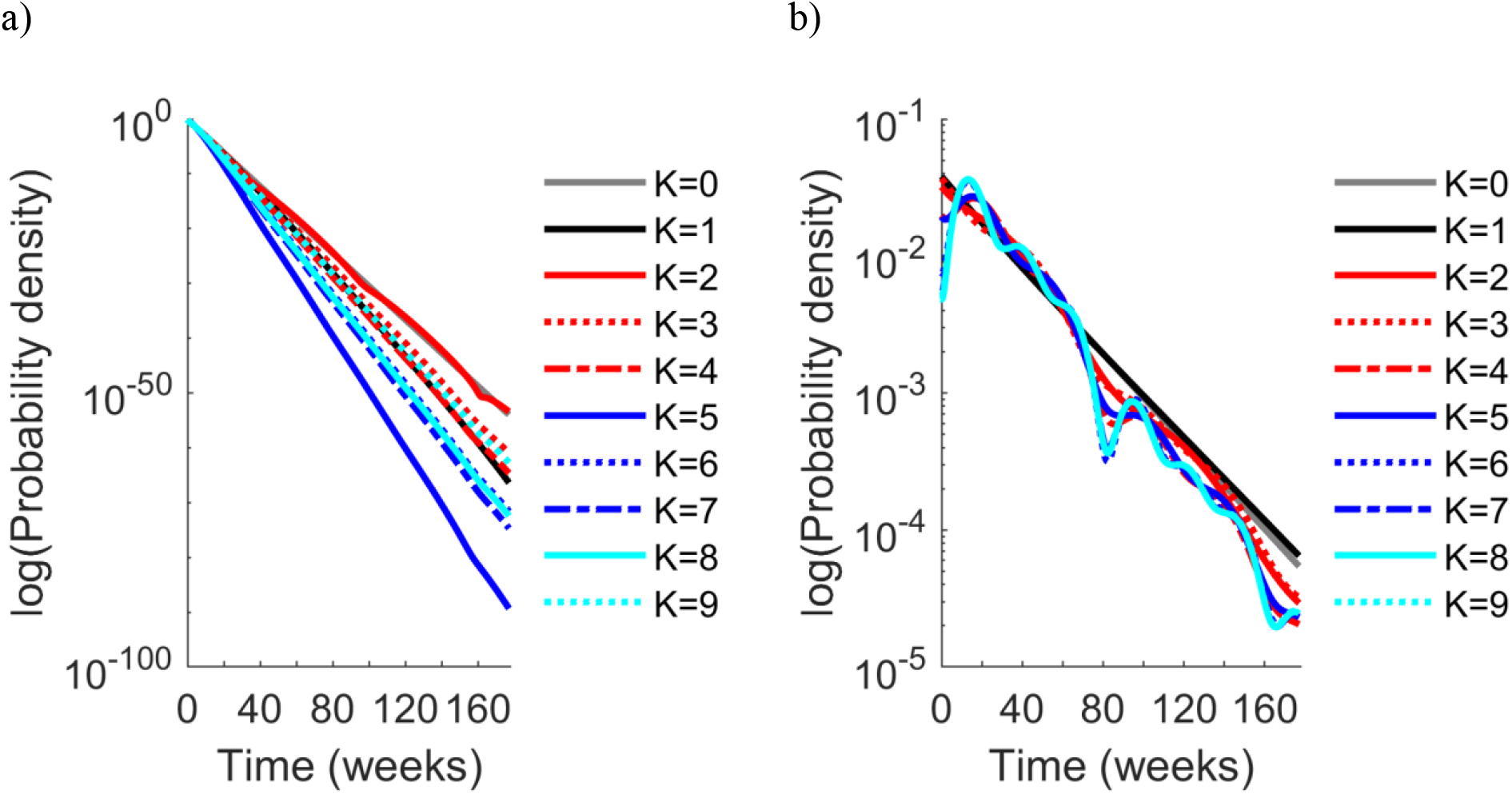
Probability density functions. The log of probability density functions for hazard rates with *K* = 0 to *K* = 9 cosine terms for a) the duration of symptomatic infection and b) the duration of asymptomatic infection.

For the optimal gSEAIR model (*K = 6*) the cosine terms of the mean residual waiting-times that are most influential correspond to a period of 82.4 days with an amplitude of 0.22 for symptomatic infection, and 817 days with an amplitude of 0.45 for asymptomatic infection (Table 2). For symptomatic infection, there were other cosine terms nearly as influential, specifically all amplitudes from *α*_*S*1_ to *α*_*S4*_where close to 0.2. For asymptomatic infection, the *α*_*A*1_term was dominant, with amplitudes *α*_*A*2_, *α*_*A4*_, and *α*_*A6*_ all about one-third of its value (Table 2).

## 4. Discussion

The analysis of our gSEAIR model illustrates it I,s an effective approach for evaluating the health burden of chlamydia in the U.S. and in assessing the potential benefits of increasing STI screening rates. The optimal gSEAIR model, according to measures such as AIC and AIC weights, was the *K = 6* scenario. This scenario predicted a greater reduction in chlamydia incidence and DALYs when increasing the annual screening rate from 35% to 40%, 50%, or 70%, at least in comparison to the traditional SEAIR model. Also, our gSEAIR models illustrated that the inclusion of time-varying average durations of infection (i.e. the mean residual waiting-times) into model dynamics typically correlated to a greater predicted health benefit from these interventions, at least in comparison to the classical SEAIR model.

As expected, the scale-up of STI screening causes a reduction in incidence and DALYs. Our findings illustrate that this reduction is comparable with other STI interventions [28], with the free distribution of condoms and diaphragms serving as a notable example [29,30]. Our predictions on this reduction are most likely conservative, as we only account for the effects of STI screening, and do not account for complementary interventions that would be deployed by health authorities, including contact tracing, partner notification [31], and the administration of suppressive therapy [32].

A particular area where the work presented here could be informative is in the rollout of Periodic Presumptive Treatment [33,34]. To elaborate, the basis of Periodic Presumptive Treatment is the systematic treatment of at-risk groups with a combination of drugs targeting prevalent (and curable) STIs, the results of which can cause reductions in STI prevalence up to 50% [34]. Thus, since our gSEAIR model provides details on multiple periods associated with transmission (i.e., the periods of the cosine terms in the mean residual waiting-times), it could help to inform on how frequent Periodic Presumptive Treatment should be deployed to maximize the health benefit of the intervention.

Although our model focuses on chlamydia transmission in the U.S., it could easily be adapted to study other STD outbreaks such as syphilis, gonorrhea, and trichomoniasis in other population demographics. Further potential generalizations and refinements include the addition of disease states, such as super-spreaders and individuals receiving treatment, the subdivision of compartments to reflect levels of at-risk behavior or age demographics, and even the generalization of the transmission rate to a pair formulation [35].

As with the majority of compartmental models, our work has several limitations. First, our model assumes a well-mixed (homogenous) population, which thereby disregards the potential impact that heterogeneity may have on the transmission cycle and intervention. Naturally, it follows that incorporating more realistic individual-level characteristics and mixing patterns would enhance the accuracy of the predictions provided. Second, the calibration of our model relies on reported chlamydia incidence from health authorities and estimates on the proportion of asymptomatic cases. Implicit in this requirement are potential biases that may arise due to myriad treatment-seeking behaviors among population groups, such as those that mistrust medical personnel, or age demographics who experience greater social stigma from disease. Finally, the model formulation imposed a parametric form of the duration of infection distribution. While the proposed distribution is flexible, as it essentially can represent any function whose Fourier cosine series converges, further empirical work is needed truly to determine the shape of the distribution.

In summary, our study provides a comprehensive evaluation of a novel form of compartmental model of chlamydia transmission. Through this model, we can better reflect recent trends in chlamydia incidence in the U.S., at least relative to traditional compartmental models, while also projecting a greater health benefit from the upscaling of STI screening interventions. By integrating the capacity to evaluate disease interventions and inform on the time periods critical to the durations of chlamydia infection, our model uniquely contributes to the wealth of knowledge needed by health officials to make informed decisions, and thereby may aid in reversing the increasing rates of STIs in the U.S.

## Data Availability

All data produced in the present work are contained in the manuscript

## Abbreviations

ODE: Ordinary Differential Equation
STD: Sexually transmitted disease
STI: Sexually transmitted infection
PID: Pelvic inflammatory disease
HIV: Human immunodeficiency virus
DALY: Disability adjusted life-years
AIC: Akaike information criterion
RSS: Residual Sum of Squares

## Acknowledgments

The authors wish to thank Dr. Emelie Kenny for constructive feedback that greatly improved the clarity of the work

SG was partially supported by the National Science Foundation Grant DMS-2052592

## Conflict of interest

All authors declare no conflict of interest in this paper

## Notes

### Competing Interest Statement

The authors have declared no competing interest.

