## Supplementary Materials for "The effect of screening on the health burden of chlamydia: An evaluation of compartmental models based on person-days of infection"

This appendix provides further details for the analysis of our compartmental model of chlamydia transmission. We outline the calculation of average duration of complications associated with chlamydial infection, give details behind our disability adjusted life-years (DALYs) calculation, and provide details on the formulation of the mean residual waiting-time used in model dynamics.

**S.1 Parameter estimation.** Here we outline the derivation of the duration of sequelae due to chlamydial infection, including the duration of moderate-to-severe pelvic inflammatory disease (PID), the duration of epididymo-orchitis, and the duration of primary and secondary infertility. We also provide details on the reproductive capabilities of both men and women and how these sequelae impact their ability to optimally reproduce within the window of average reproductive capability.

**S.1.1 The transmission rate of chlamydia.** From our model, we estimate the average duration of moderate PID, $\lambda_M$, and severe PID, $\lambda_D$, due to infection

Table S.1. Estimated transmission rates for SEAIR and gSEAIR models

| | $K$ | | | | | | | | | |
|---|---|---|---|---|---|---|---|---|---|---|
| | 0 | 1 | 2 | 3 | 4 | 5 | 6 | 7 | 8 | 9 |
| $\beta$ | 0.0524 | 0.0498 | 0.0565 | 0.0555 | 0.0615 | 0.0607 | 0.0611 | 0.0616 | 0.0617 | 0.0614 |

**S.1.2 The duration of moderate and severe pelvic inflammatory disease.** From our model, we estimate the average duration of moderate PID, $\lambda_M$, and severe PID, $\lambda_D$, due to infection in women by

$$\lambda_M = \gamma_{RCF} - \varrho_F - \vartheta_{FP} = \lambda_D \quad \text{(S1)}$$

where $\gamma_{RCF}$ represents the number of years a female is able to reproduce, $\varrho_F$ represents the average time that a female is able to reproduce before potentially contracting chlamydial infection, and $\vartheta_{FP}$ represents the years lost in reproductivity due to PID (see Table 1 for details).

We also assume that the time when a female begins being fertile is at the onset of menarche, which on average begins at 12.5 years [1]. From the literature, the average length of reproductive capability in females is 39 years [2]. It also follows that there are 8.5 years of reproductive capability in women before the average age that a female contracts chlamydial infection, which is at 21 years, assuming uniformity across the United States population [3]. Thus, it follows that

$$\gamma_{RCF} \approx 39 \text{ years and } \varrho_F \approx 8.5 \text{ years.}$$

To determine $\vartheta_{FP}$, we estimate the average time for a woman asymptomatically infected with chlamydia to develop PID. As 1 in 10 women infected with chlamydia develops PID within a year [4], assuming an exponential distribution, we have that

$$e^{-1.00x} = 0.9.$$

It follows that the rate of PID is

$$x = 0.10536/\text{year}$$

and consequently,

$$\vartheta_{FP} = \int_0^{\infty} e^{-0.10536t} dt \approx 9.49 \text{ years.}$$

Substituting into (S1) yields

$$\lambda_M = \lambda_D = 21.01 \text{ years.}$$

Furthermore, because PID is most common in individuals aged 15-24, we assume the average age of PID onset is $\lambda_{PID} = 19.5$. Thus, the years of life lost from death due to PID is

$$\lambda_{Death} = L - \lambda_{PID} - \lambda_D = 36.5 \; years,$$

where $L = 77$ years is the average lifespan in the U.S. [5].

**S.1.3 The proportion of cases classified as severe PID**. We estimate the proportion of moderate and severe PID cases based on published data, which estimates that 15%-20% of PID cases are severe [6]. Thus, we assume

$$\rho_D = \frac{0.15 + 0.2}{2} = 0.175,$$

The proportion of moderate PID cases are taken as $1 - \rho_D$.

**S.1.4 The duration of epididymo-orchitis.** Epididymo-orchitis is a common complication of chlamydial infection in males [7], which causes infertility [8].

To determine the average duration of complications associated with epididymo-orchitis, $\lambda_E$, we assume that

$$\lambda_E = \gamma_{RCM} - \varrho_M - \vartheta_{ME} \qquad \text{(S2)}$$

where $\gamma_{RCM}$ represents the number of years a male is capable of reproducing, $\varrho_M$ represents the average age that a male contracts chlamydial infection, and $\vartheta_{ME}$ represents the years lost in reproductivity due to epididymo-orchitis infection (see Table 1 for more details).

We define the ability of a male to reproduce as the time when sperm are most motile and representative of optimal fertility. We assume this time begins at the onset of spermarche, which on average begins at 13.4 years [9] and continues to age 55, which is when motility counts begin to diminish [10]. It follows that

$$\gamma_{RCM} \approx 41.6 \text{ years.}$$

In addition, we base $\varrho_M$ on the highest prevalence between 20 to 24 years old [11]. Taking the average, it follows that the average age a male contracts chlamydial infection is 22 years. Thus, it follows that

$$\varrho_M \approx 8.6 \text{ years } [11].$$

To determine $\vartheta_{ME}$, we estimate the average time for a male asymptomatically infected with chlamydia to develop epididymo-orchitis. Based on the literature, 12.3% of infected men develop epididymo-orchitis within a year [12]. Assuming an exponential distribution, this would imply that

$$e^{-1.00x} = 0.877$$

Thus, it follows that the rate of epididymo-orchitis is

$$x = 0.13124 \text{ /year.}$$

To determine $\vartheta_{ME}$, it follows that

$$\vartheta_{ME} = \int_0^\infty e^{-0.13124t} dt \approx 7.62 \text{ years.}$$

Substituting into (S2) yields

$$\lambda_E \approx 25.38 \text{ years.}$$

**S.2. Disability adjusted life-years.** We quantify the health burden caused by chlamydia using the health metric Disability adjusted life-years (DALYs). Specifically, we calculate time-discounted DALYs through years lived with disability (YLD) and the years of life lost (YLL):

$$YLD = \big((1-\phi)\rho_I\omega_I\lambda_I + \phi\rho_A\omega_A\lambda_A + \rho_M(1-\rho_D)\omega_M\lambda_M + \tau\rho_M\rho_D(1-\rho_{Death})\omega_D\lambda_D + (1-\tau)\rho_E\omega_E\lambda_E\big) \cdot \int_0^5 \frac{\beta(I_A+I_S)S}{N} e^{-\frac{0.03}{365}t} dt$$

and

$$YLL = \tau\rho_M\rho_D\rho_{Death}\lambda_{Death} \cdot \int_0^5 \frac{\beta(I_A+I_S)S}{N} e^{-\frac{0.03}{365}t} dt.$$

Here, $\rho_k$, $\omega_k$, and $\lambda_k$ are the proportion, DALY weight, and average duration of the $k^{th}$ outcome associated to chlamydia infection, as outlined in Table 1, and $\tau$ is the proportion of chlamydial infections held by women. It follows that the total DALYs for the $i^{th}$ scenario is

$$\text{Total DALYs}_i = YLL + YLD.$$

The DALYs averted from the $i^{th}$ scenario are then determined by subtracting the total DALYS of a scenario from the baseline:

$$\text{DALYs averted}_i = \text{Total DALYs}_{Baseline} - \text{Total DALYs}_i.$$

**S.3 A class of distributions with periodic hazard rate.** The distribution for our model's infectious period is based on a extending the Probability density function (PDF) [13]

$$f(t) = \frac{1}{\frac{1}{\rho} - \frac{\rho\alpha}{\rho^2 + w^2}} (1 - \alpha\cos(wt))e^{-\rho t},$$

where $\alpha \in (-1,1)$, $\rho > 0$, and $w \geq 0$, to that of a Fourier cosine series:

$$f(t) = \frac{1}{\frac{1}{\rho} - \sum_{i=1}^{N} \frac{\rho\alpha_i}{\rho^2 + w_i^2}} \left(1 - \sum_{i=1}^{N} \alpha_i \cos(w_i t)\right) e^{-\rho t},$$

where $\rho > 0, a_i \in (-1,1)$, and $\sum_{i=1}^{N} |a_i| < 1$.

Our distribution exhibits several convenient properties, including a closed-form Cumulative distribution function (CDF)

$$F(t) = \frac{1}{\frac{1}{\rho} - \sum_{i=1}^{N} \frac{\rho\alpha_i}{\rho^2 + w_i^2}} \left(\frac{1}{\rho} - \frac{1}{\rho} e^{-\rho t} - \sum_{i=1}^{N} \frac{\alpha_i}{\rho^2 + w_i^2} (\rho - (\rho \cos(w_i t) - w_i \sin(w_i t)) e^{-\rho t})\right).$$

It also has a closed-form (and periodic) mean residual waiting-time

$$m(t) = \frac{1 - \rho^2 \sum_{i=1}^{K} a_i \frac{(\rho^2 - w_i^2)\cos(w_i t) - 2\rho w_i \sin(w_i t)}{(\rho^2 + w_i^2)^2}}{\rho - \rho^2 \sum_{i=1}^{K} \frac{a_i\big(\rho \cos(w_i t) - w_{ji} \sin(w_i t)\big)}{\rho^2 + w_i^2}}$$

and hazard rate

$$\eta(t) = \frac{\rho(1 - \sum_{i=1}^{K} a_i \cos(w_i t))}{1 - \rho \sum_{i=1}^{K} \frac{a_i(\rho \cos(w_i t) - w_i \sin(w_i t))}{\rho^2 + w_i^2}}.$$

On account that $\mu = m(0)$, it follows that the average infectious period for our distribution is

$$\mu = m(0) = \frac{1 - \rho^2 \sum_{i=1}^{K} a_i \frac{(\rho^2 - w_i^2)}{(\rho^2 + w_i^2)^2}}{\rho - \rho^2 \sum_{i=1}^{K} a_i \frac{\rho}{\rho^2 + w_i^2}},$$

and that the variance can be computed using

$$\sigma^2 = \int_0^\infty 2t\big(1 - F(t)\big)dt - \mu^2 = \frac{2\left(1 - \rho^4 \sum_{i=1}^{N} a_i \frac{\rho^2 - 3w_i^2}{(\rho^2 + w_i^2)^3}\right)}{\rho^2\left(1 - \rho \sum_{i=1}^{N} a_i \frac{\rho}{\rho^2 + w_i^2}\right)} - \mu^2.$$
